# COVID-19 Convalescent Plasma Outpatient Therapy to Prevent Outpatient Hospitalization: A Meta-analysis of Individual Participant Data From Five Randomized Trials

**DOI:** 10.1101/2022.12.16.22283585

**Authors:** Adam C. Levine, Yuriko Fukuta, Moises A. Huaman, Jiangda Ou, Barry R. Meisenberg, Bela Patel, James H. Paxton, Daniel F. Hanley, Bart JA Rijnders, Arvind Gharbharan, Casper Rokx, Jaap Jan Zwaginga, Andrea Alemany, Oriol Mitjà, Dan Ouchi, Pere Millat-Martinez, Valerie Durkalski-Mauldin, Frederick K. Korley, Larry J. Dumont, Clifton W. Callaway, Romina Libster, Gonzalo Perez Marc, Diego Wappner, Ignacio Esteban, Fernando Polack, David J. Sullivan

## Abstract

**Background:** Monoclonal antibody and antiviral treatments for COVID-19 disease remain largely unavailable worldwide, and existing monoclonal antibodies may be less active against circulating omicron variants. Although treatment with COVID-19 convalescent plasma (CCP) is promising, randomized clinical trials (RCTs) among outpatients have shown mixed results.

**Methods:** We conducted an individual participant data meta-analysis from all outpatient CCP RCTs to assess the overall risk reduction for all-cause hospitalizations by day 28 in all participants who had transfusion initiated. Relevant trials were identified by searching MEDLINE, Embase, MedRxiv, WHO, Cochrane Library, and Web of Science from January 2020 to September 2022.

**Results:** Five included studies from four countries enrolled and transfused 2,620 adult patients. Comorbidities were present in 1,795 (69%). The anti-Spike or virus neutralizing antibody titer range across all trials was broad. 160 (12.2%) of 1315 control patients were hospitalized, versus 111 (8.5%) of 1305 CCP-treated patients, yielding a 3.7% (95%CI: 1.3%-6.0%; p=.001) ARR and 30.1% RRR for all-cause hospitalization. The effect size was greatest in those with both early transfusion and high titer with a 7.6% ARR (95%CI: 4.0%-11.1%; p=.0001) accompanied by at 51.4% RRR. No significant reduction in hospitalization was seen with treatment > 5 days after symptom onset or in those receiving CCP with antibody titers below the median titer.

**Conclusions:** Among outpatients with COVID-19, treatment with CCP reduced the rate of all-cause hospitalization. CCP may be most effective when given within 5 days of symptom onset and when antibody titer is higher.

**Key Points:** While the outpatient COVID-19 randomized controlled trial meta-analysis indicated heterogeneity in participant risk factors and convalescent plasma, the combined CCP efficacy for reducing hospitalization was significant, improving with transfusion within 5 days of symptom onset and high antibody neutralization levels.

## Introduction

Globally, the COVID-19 pandemic has claimed more than 18 million lives, including over 1 million deaths in the United States (US) alone.^1^ Despite widespread vaccination in high- and middle-income countries, new variant outbreaks continue to fuel economic disruptions and increased hospitalization.^2^ Novel vaccines and treatments against SARS-CoV-2 have been developed, tested, and deployed in record time, yet most arrived too late to benefit the millions of people who died in the pandemic’s first year.^1^ Three years into the COVID-19 pandemic, it remains unclear how we can respond faster and more effectively to the next pandemic.^3, 4^

Antibodies to the SARS-CoV-2 virus, whether induced by vaccination or infused as monoclonal antibodies (mAbs) or polyclonal convalescent plasma, have been shown to reduce the risk of COVID-19 related hospitalization and death, but only convalescent plasma is likely to be both available and affordable for the majority of the world’s population in the early days of the next viral pandemic.^5^ COVID-19 convalescent plasma (CCP) was first administered to a hospitalized patient on March 28, 2020,^6^ just two weeks after the World Health Organization declared a pandemic. Meanwhile, mAbs to prevent hospitalization^7, 8^ and vaccines^9, 10^ to prevent symptomatic infection, hospitalization, or death were not available until December 2020. By that time, more than 79 million cases of COVID-19 and 1.7 million deaths had been reported worldwide^11^. Effective oral drug therapy for outpatient use was not available until a year later, in December 2021.^12^ While a safe and effective oral agent against SARS-CoV-2 is the ideal solution to prevent COVID-19 hospitalization, this solution remains unavailable to many patients worldwide due to high costs,^13, 14^ and its effectiveness could be threatened at any time by new unsusceptible variants.

Of the two remaining mAbs effective against omicron variants, one has been shown to have reduced in vitro activity and has only been approved for prophylaxis, not treatment.^15-17^ Furthermore, escape mutations in the spike protein leading to acquired resistance during treatment with a single mAb have been repeatedly described in immunocompromised patients.^18, 19^ The rapid rise of variants with mutations in the spike protein has created a dilemma in mAb development, as pharmaceutical companies must weigh the high cost of their development against the short-lived utility of these agents^20^. Now that mAbs are proving less effective against mutant strains, CCP remains an important therapeutic option, especially for severely immunocompromised and other high-risk patients.^21, 22^

Most randomized controlled trials (RCTs) of CCP were conducted in patients already hospitalized with COVID-19, largely due to the convenience of conducting research in this population. Later in the pandemic, RCTs of CCP targeting outpatients were designed to determine whether early treatment could prevent hospitalization. Our objective in this study was to conduct an individual patient meta-analysis of all available RCTs of CCP in adult COVID-19 outpatients to determine whether early CCP therapy can reduce the risk of hospitalization.

## METHODS

This study followed the guidelines provided in the Preferred Reporting Items for Systematic Reviews and Meta-Analyses (PRISMA) 2020 statement.^23^

### Objectives

This review aimed to find, assess, and synthesize all RCTs that assessed the efficacy of CCP in preventing all-cause hospitalization among outpatients with confirmed SARS-CoV-2 infection.

### Eligibility, Search Strategy, RCT Selection, Data Extraction and Quality

Our PICO (population, intervention, comparator, and outcome) therefore included the following: population = adult (≥18 years) COVID-19 outpatients, regardless of risk factors; intervention = intravenous COVID-19 convalescent plasma transfusion, regardless of antibody titer; comparators = control (e.g., non-convalescent plasma, normal saline, multi-vitamin); outcome = all-cause hospitalization within 28 days of transfusion. For one study in Argentina, patients meeting prespecified hypoxic respiratory criteria were sometimes admitted to a specific unit within their long-term care facilities, which provided hospital-level care, to avoid overcrowding hospitals. For purposes of trial eligibility, we considered these admissions to be hospitalizations. Only English-language documents were reviewed.

A literature search was performed independently by two authors (YF, DJS). The MEDLINE, Embase, MedRxiv, Cochrane Library, WHO COVID-19 Research Database, and Web of Science were searched for all RCTs as of 30 September 2022. Search strategies were designed with terms related to CCP and COVID-19 (supplementary Figure 1). All RCTs were included that met the eligibility criteria above. We contacted the corresponding authors for each of the included trials and asked them to contribute data and serve as co-authors for the prepared manuscript.

The investigators for each RCT provided the following data elements: trial design characteristics, descriptions of the intervention and control groups, baseline characteristics of the patients (including underlying comorbidities and days after symptom onset), CCP characteristics (e.g., antibody titers; etc.), hospitalizations, enrollment period, target enrollment, number of enrollments, number of transfusions, and trial locations. Data not provided in the published reports were collected from the authors.

A risk of bias assessment for each selected trial was performed by COVID-19 Network Meta-Analysis (NMA).^24, 25^

### Statistical Method

Primary and secondary analyses were done in the modified intention-to-treat population including all randomized participants who received the intervention (either CCP or control). The primary outcome used for analysis was all-cause hospitalization within 28 days of transfusion, and the secondary outcome was all-cause hospitalization minus those patients admitted to hospitals within 24 hours of transfusion. Two subgroup analyses were performed: 1) the effect on hospital admission for patients with ≤5 versus >5 days of symptoms at the time of intervention; and, 2) the effect on patients receiving CCP with antibody titers above the median SARS-CoV2 antibody titer value for each individual RCT versus those receiving CCP not above the median.

Descriptive analysis included the country in which the study was conducted, patient demographics, days since symptom onset, plasma donor antibody levels, and high-risk comorbidities. Box plots were used for visualization and comparison of viral neutralization among studies. Treatment effect was determined using the absolute risk reduction (ARR), relative risk reduction (RRR), and number needed to treat (NNT). Odds ratio (OR), 95% CI, weight of each study (inverse of the variances), heterogeneity (*I*^*2*^), between-study variance (*τ*^*2*^) and significance levels were estimated using random effect models and displayed in forest plots. A funnel plot was used to estimate the risk of publication bias. The significance level for analyses was set at 0.05, and statistical tests were two-tailed. All the data manipulation and analyses were performed using Excel, R (version 4.2.0, R Foundation, Vienna, Austria) and its statistical packages “meta” (version 6.0-0) and “metafor” (version 3.8-1).

## RESULTS

### Trial population

A total of 617 studies were identified by our primary search strategy. After screening and exclusion of ineligible studies, five RCTs were included (Figure 1). Of these, two were conducted in the United States^26, 27^, two in Europe^28, 29^ and one in Argentina.^30^ All the trials were stopped early: one due to slow recruitment as COVID-19 cases in the trial region decreased considerably,^30^ three due to rapid uptake of vaccination resulting in substantial reduction in hospital admission rates,^26, 28, 29^ and one due to a finding of futility to detect the planned difference after the second planned interim analysis of the primary outcome alysis.^27^

**Figure 1.**
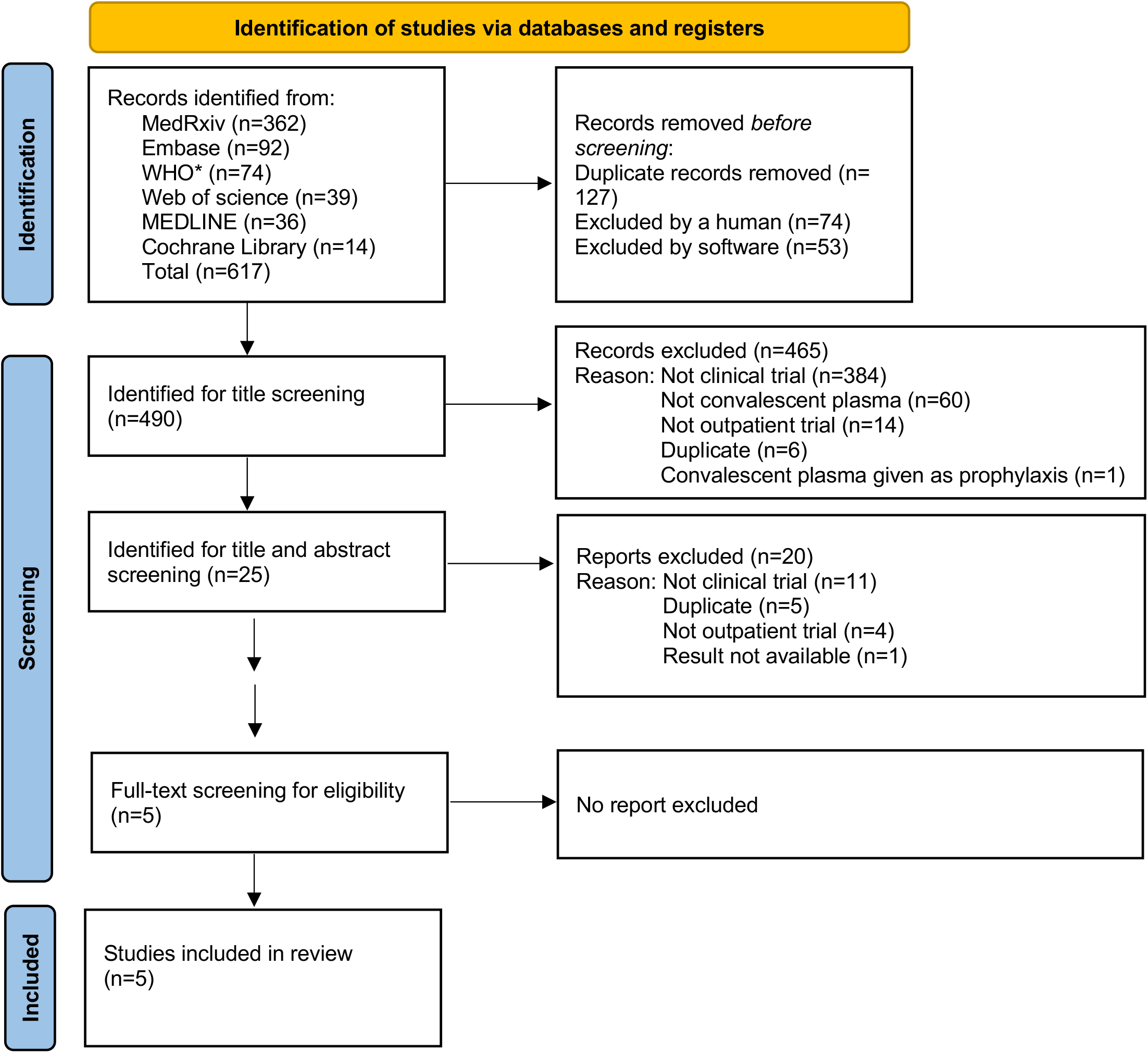
PRISMA chart *WHO COVID-19 global literature on coronavirus disease

The five included RCTs recruited patients from June 2020 to October 2021.^26-30^ Trial enrollment totals ranged from 154 to 1,181, yielding a pooled analysis sample of 2,620 patients with early COVID-19 transfused with either CCP or control. These trials were varied in terms of their demographic and clinical profiles, including median age, sex distribution, and the prevalence of major risk factors for COVID-19-related hospitalization (Table 1). Studies also varied somewhat in the timing of the intervention, although 1,562 patients (60%) were transfused within five days of symptom onset. Overall, only 159 (6%) of all patients were fully vaccinated against COVID-19. We found that the risk of bias was low for the five RCTs, (Supplementary Table 1). Funnel plot analysis shows a low risk of publication bias (Supplementary Figure 2).

**Table 1.** Trial characteristics

|  | **CSSC-004** | **CCP**<br>**Argentina** | **CONV-**<br>**ERT** | **C3PO** | **CoV-Early** | **total**<br>**all 5** |
| --- | --- | --- | --- | --- | --- | --- |
| control arm | plasma | saline | saline | saline/multi -<br>vitamin | plasma |  |
| enrollment period | June 2020<br>to Oct 2021 | June 2020<br>to Oct<br>2020 | Nov 2020<br>to July<br>2021 | Aug 2020 to<br>Feb 2021 | Nov 2020<br>to July 2021 |  |
| Trial duration (months) | 16 | 5 | 9 | 7 | 9 | 46 |
| variants | 614G,<br>alpha, beta,<br>delta | WA-1,<br>D614G | D614G,<br>alpha | D614G | D614G,<br>alpha |  |
| geography | USA | Argentina | Spain | USA | Netherlands |  |
| target enrollment | 1344 | 210 | 474 | 900 | 690 | 3318 |
| enrolled | 1225 | 160 | 376 | 511 | 421 | 2693 |
| mITT | 1181 | 154 | 369 | 500 | 416 | 2620 |
| median age (range) | 43 (18-85) | 77 (65-90+) | 56 (IQR<br>52–62) | 54 (18-93) | 60 (IQR 55-65) |  |
| 1+ medical high risk<br>condition for COVID-19<br>progression (%) | 470 (40) | 131 (82) | 278 (74) | 511 (100) | 416 (100) | 1806<br>(68.6) |
| symptoms <= 5 days (%) | 517 (44) | 154 (100) | 283 (77) | 389 (78) | 226 (54) | 1569<br>(60) |
| symptoms <= 3 days (%) | 168 (14) | 154 (100) | 101 (27) | 240 (48) | 52 (13) | 715<br>(27) |
| median/mean duration<br>symptoms | 6 | 3 | 4.4 | 4 | 5 |  |
| total female (%) | 675 (57) | 98 (64) | 169 (46) | 265 (53) | 93 (22) | 1300<br>(50) |
| age over 50 (%) | 411 (35) | 154 (100) | 368 (100) | 310 (61) | 414 (100) | 1657<br>(63) |
| age over 65 (%) | 80 (7) | 154 (100) | 73 (20) | 95 (19) | 113 (27) | 515<br>(20) |
| diabetes (%) | 99 (8) | 35 (23) | 39 (10) | 142 (28) | 29 (7) | 344<br>(13) |
| hypertension (%) | 276 (23) | 110 (71) | 244 (66) | 216 (42) | not reported | 846<br>(38) † |
| obesity or BMI >30 (%) | 444 (38) | 11 (7) | 95 (26) | 302 (60) | 126 (30) | 978<br>(37) |

†Only included 4 reported studies.
Abbreviations. MVC=multi-vitamin concentrate; BMI=body mass index; mITT=modified intention to treat

### Convalescent plasma

The included studies used a variety of assays to qualify and characterize the CCP transfused in study subjects (Supplementary Table 2). Unfortunately, there was insufficient residual donor plasma samples available to compare neutralization titers across the different studies using the same assay. Two studies qualified units with 50% viral neutralization dilutional plasma titers greater than 1:160. Two studies qualified with dilutional antibody binding greater than 1000 or 320, while the last measured Euroimmun IgG over 6.0 AU. Viral neutralization indices, depicted in Supplementary Figure 3, show that the CSSC-004 and CCP-Argentina had slightly lower viral neutralization metrics, albeit a different viral neutralization assay than C3PO, CoV-Early and CONV-ERT.

### Primary outcome: Hospitalization

A modified intention to treat analysis (Table 2) was performed on patients who received either CCP or control, excluding 6 subjects (4%) from the original study population of the CCP-Argentina and 11 (2%) from the C3PO trial who did not receive the treatment to which they were randomized for the primary outcome of all-cause hospitalization. CSSC-004 added 7 all cause hospitalizations (4 CCP and 3 control plasma) above reported COVID-19 related hospitalizations and C3PO added two participants hospitalized after day 15 before day 28. Overall, 160 (12.2%) subjects in the control group were hospitalized, compared to 111 (8.5%) in the CCP treatment group, yielding an ARR of 3.7% (95%CI: 1.3%-6.0%) and RRR of 30.1% (95%CI: 12.0%-44.4%) for all-cause hospitalization (Table 2). The OR for hospitalization was 0.64 (95%CI: 0.45-0.92) in the pooled meta-analysis, and trial heterogeneity was moderate, with an I^2^ of 42% (Figure 2). A secondary analysis was conducted excluding those patients admitted to the hospital within 24-hours of CCP (25 patients) or control (13 patients) transfusion, yielding an ARR of 4.4% (95%CI: 2.2%-6.6%) and RRR of 39.2% (95%CI: 21.7%-52.8%). The OR for hospitalization was 0.58 (95%CI: 0.41-0.82), and trial heterogeneity was low in this secondary analysis, with an I^2^ of 31% (Figure 2).

**Table 2.**
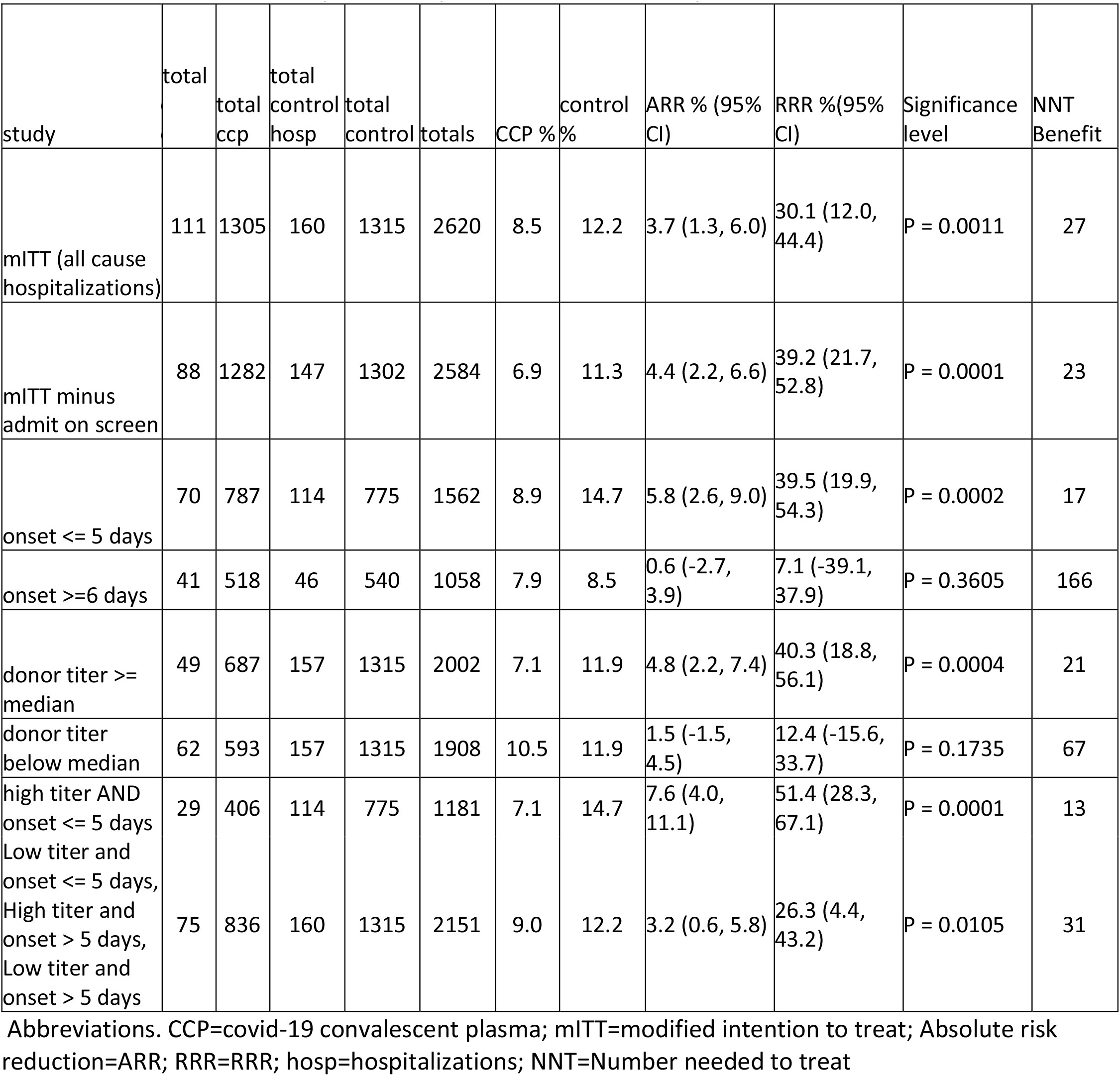
Overall numbers and percent of pooled numbers for hospitalization and totals

**Figure 2.**
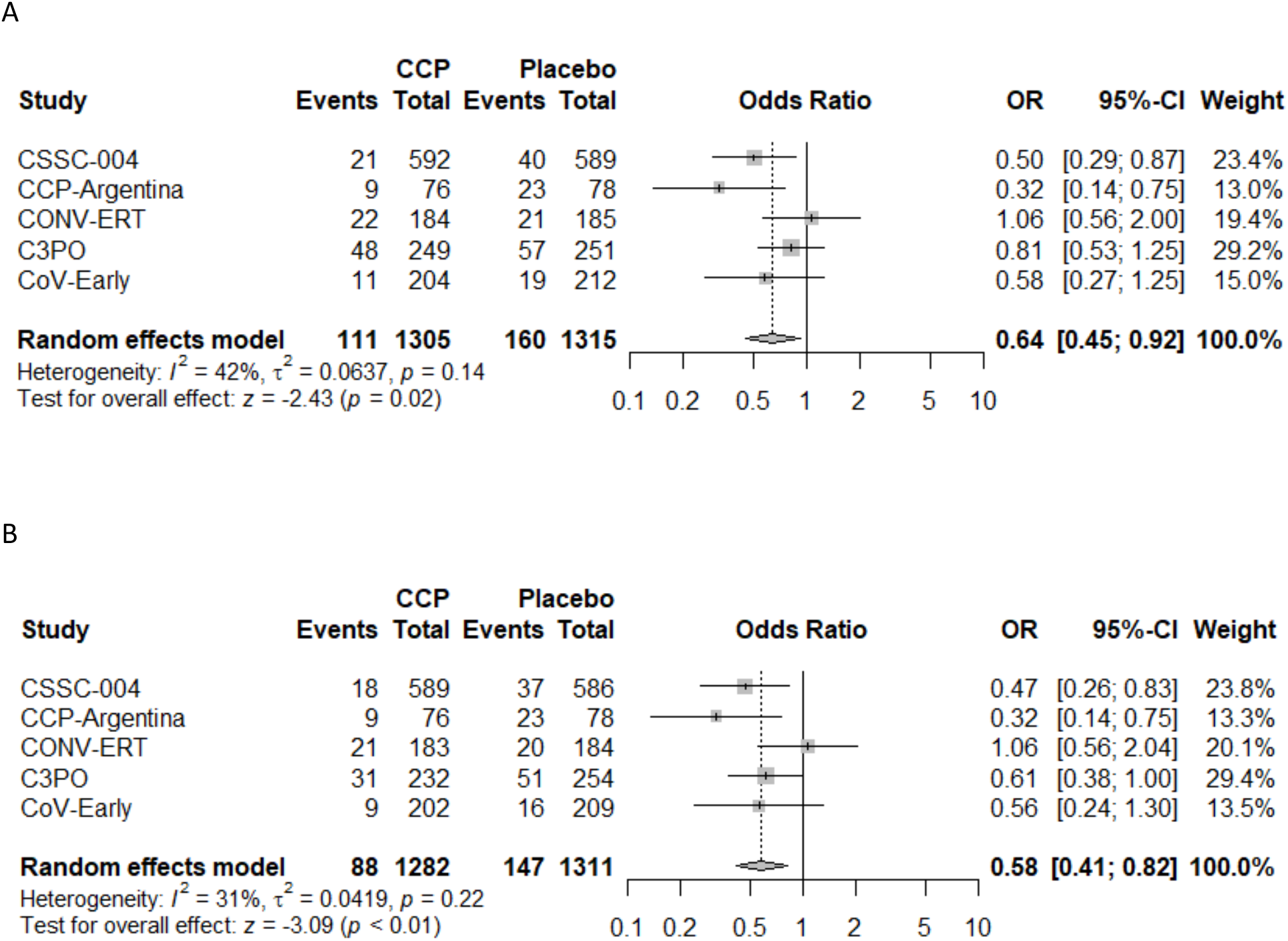
Forest plot of A) modified Intention to Treat Analysis and B) of modified Intention to Treat Analysis excluding same day hospital admissions Abbreviations. CCP=COVID-19 convalescent plasma; OR=odds ratio; CI=confidence interval

### Subgroup Analyses

Subgroup analyses were performed based upon the timing of CCP transfusion and the SARS-CoV-2 antibody titer level in transfused CCP units. For subjects transfused within 5 days of symptom onset, pooled analysis amongst all five studies indicated a 5.8% (95%CI: 2.6%-9.0%) ARR and 39.5% (95%CI: 19.9%-54.3%) RRR in hospitalizations when compared to control (Table 2 and Figure 3). Study subjects transfused with high-antibody titer CCP (defined as > than the median neutralization titer for each individual study) had an ARR of 4.8% (95%CI: 2.2%-7.4%) and RRR of 40.3% (95%CI: 18.8%-56.1%) in hospitalization when compared to subjects given the control (Table 2 and Figure 4). Subjects transfused after 6 days of symptoms or with low antibody titer CCP did not show a significant decrease in hospitalization when compared with control (Table 2). The risk reduction in patients receiving high antibody titer CCP AND within 5 days of symptom onset was higher for the combined studies at 7.6% (95%CI: 4.0%-11.1%) ARR and 51.7% (95%CI: 28.3%-67.1%) RRR (Table 2 and Figure 5).

**Figure 3.**
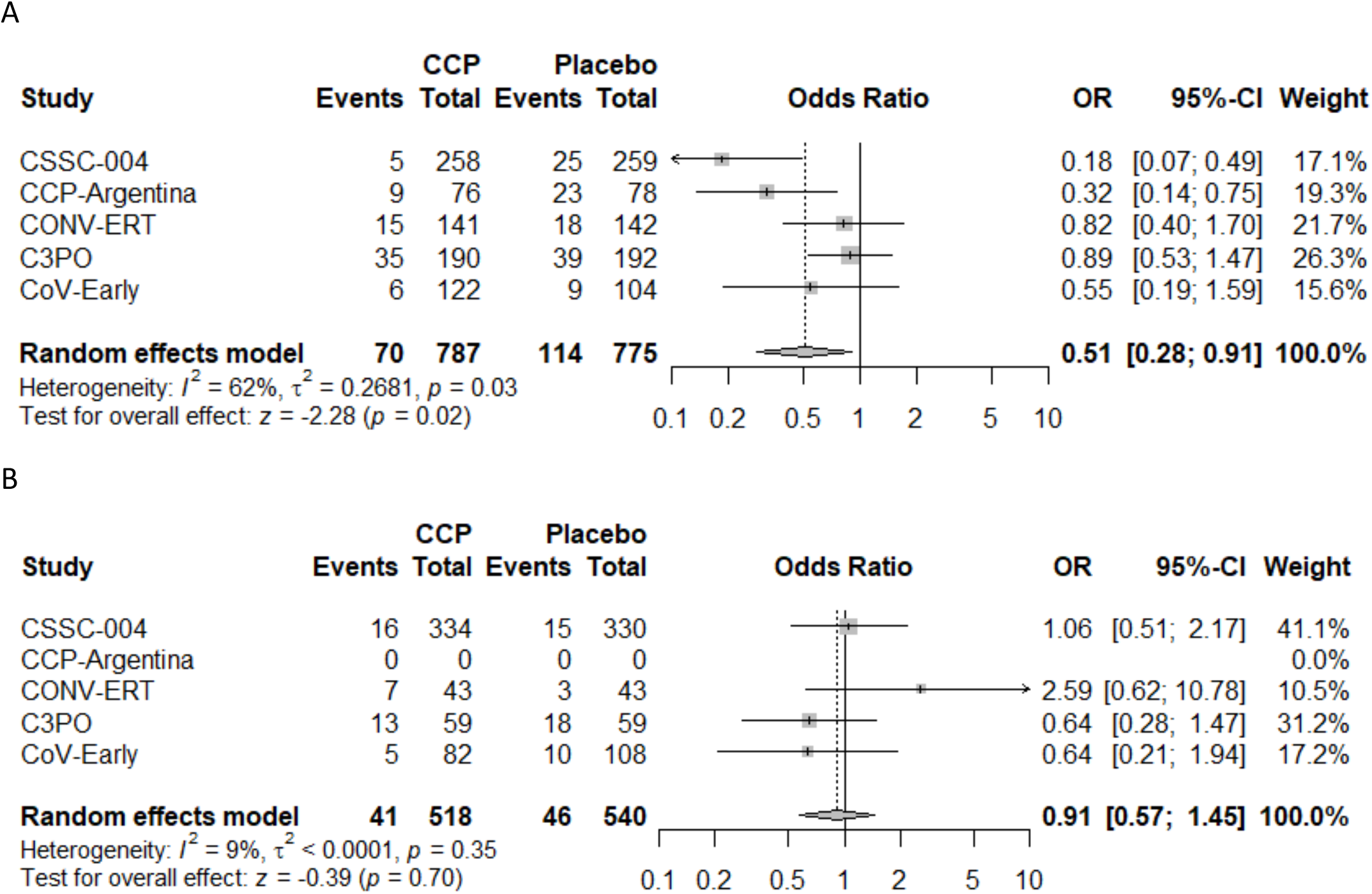
Forest plots of transfusion A) within 5 days or B) greater than 5 days A Abbreviations. CCP=COVID-19 convalescent plasma; OR=odds ratio; CI=confidence interval

**Figure 4.**
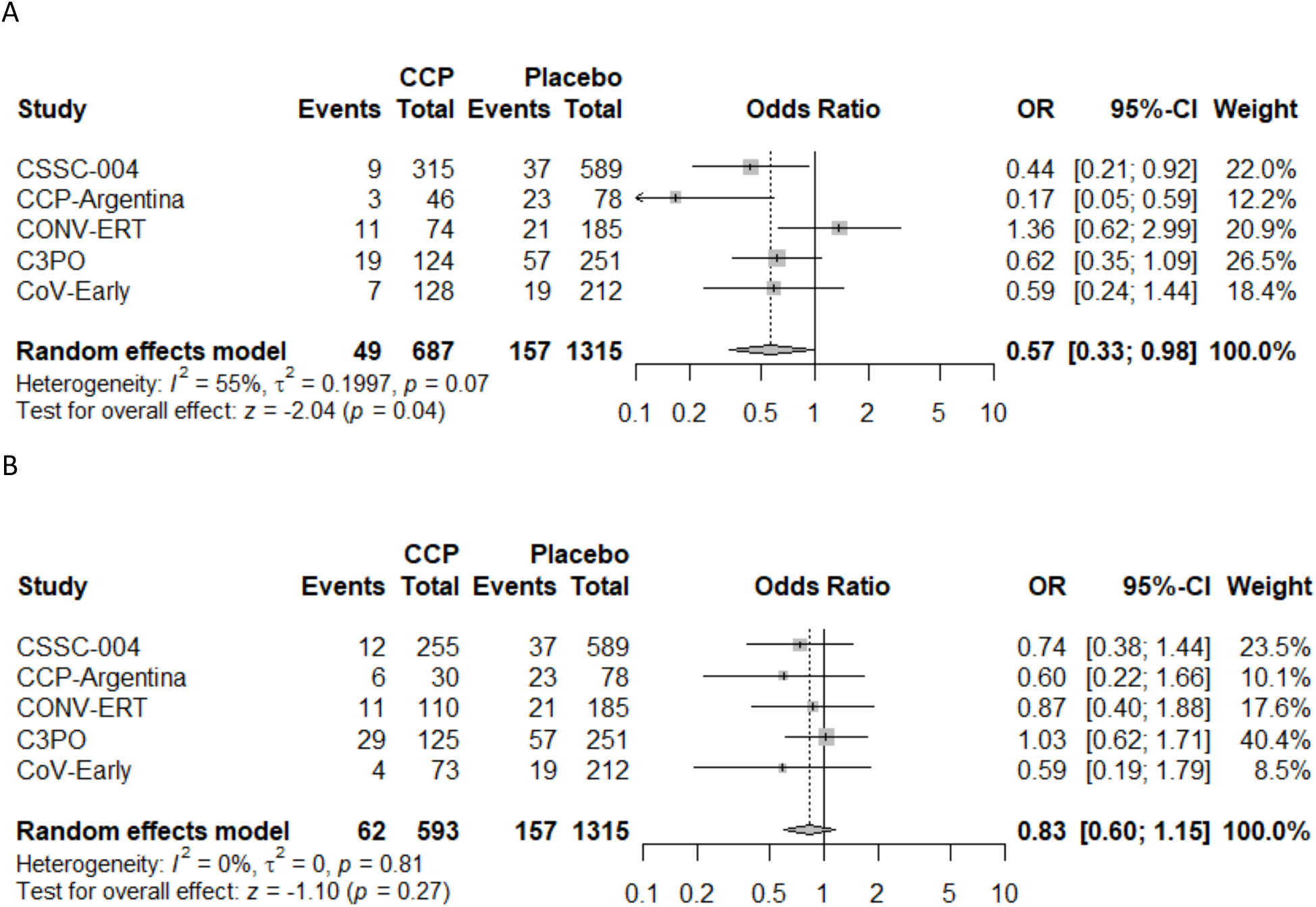
Forest plots plasma donor antibody levels A) at or above median titer or B) less than median titer Abbreviations. CCP=COVID-19 convalescent plasma; OR=odds ratio; CI=confidence interval

**Figure 5.**
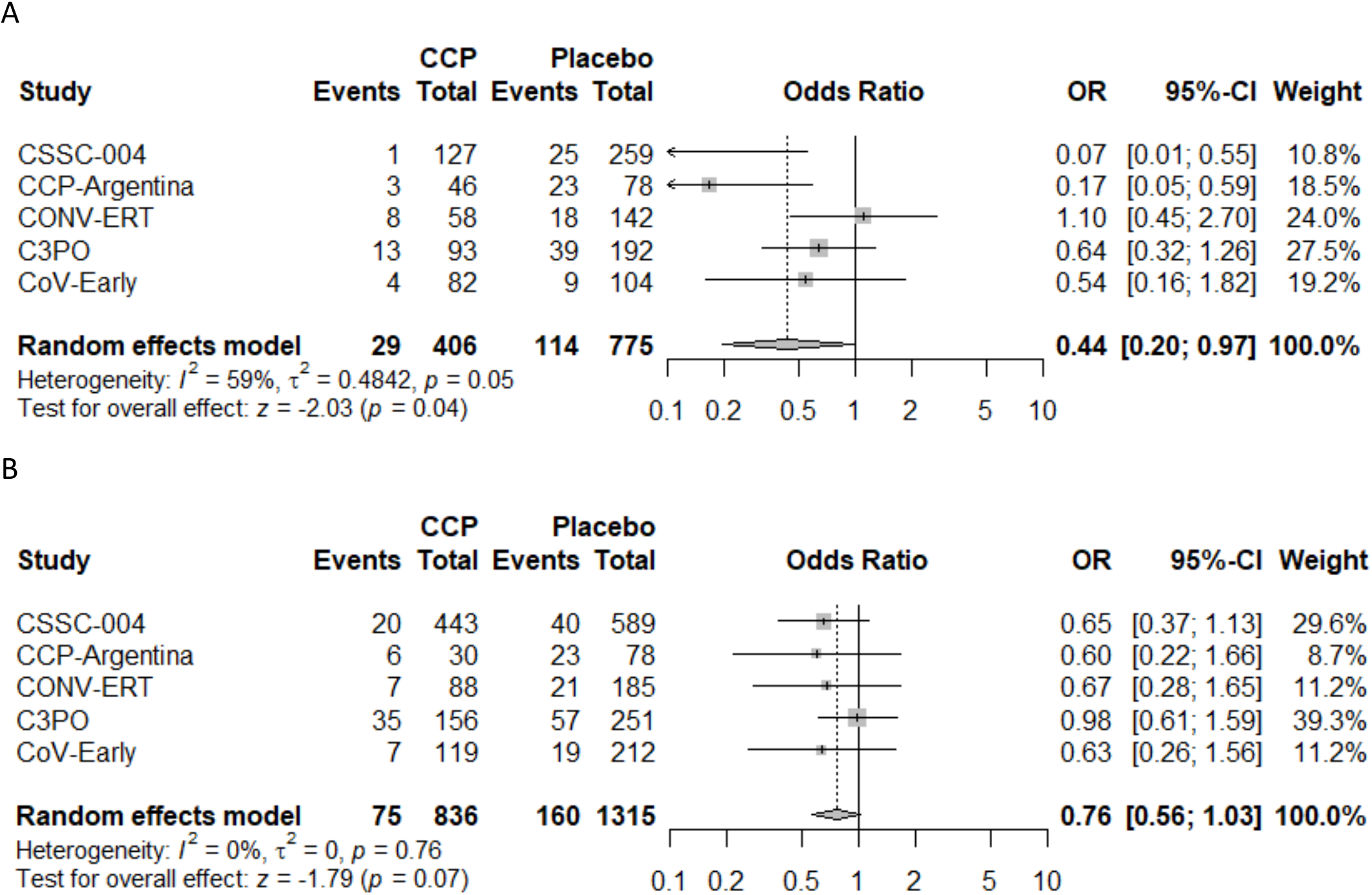
Forest plots plasma donor antibody levels and early treatment A) at or above median titer AND transfusion within 5 days or B) Total of (Low titer and onset<= 5 days, High titer and onset over 5 days, Low titer and onset over 5 days Abbreviations. CCP=COVID-19 convalescent plasma; OR=odds ratio; CI=confidence interval

### Safety

Due to small numbers, we did not combine severe adverse events in a meta-analysis; however, they were collected for each trial. In CSSC-004, one subject experienced a transfusion reaction that required cessation of the transfusion. Another transfusion was stopped due to the appearance of mild hives at the patient’s request.^26^ The CCP-Argentina trial did not report any instances of volume overload, allergic reactions, or vasovagal syndromes, but did report one case of thrombophlebitis in the control arm. The C3PO authors noted three serious transfusion reactions in the CCP arm resulting in steroid or epinephrine administration or admission to the hospital.^27^ The CONV-ERT team communicated treatment-related events^29^ with no Transfusion related acute lung injury (TRALI), transfusion associated circulatory overload (TACO), or anaphylaxis in the CCP arm, but vasovagal reactions in 3 subjects transfused with control and 1 subject transfused with CCP, and mild allergic reactions in 12 (6.4%) of 188 subjects transfused with CCP. One subject developed a pulmonary embolism 7 days after CCP transfusion. The CoV-Early investigators reported three serious adverse events possibly related to plasma transfusion (all with non-convalescent plasma). Two developed an anaphylactic reaction shortly after receiving plasma for which no hospital admission was required and one patient developed generalized urticaria and was hospitalized.

## Discussion

This meta-analysis of all available RCTs found that early outpatient therapy with CCP in adult patients with COVID-19 was associated with an 30% all cause hospitalization RRR for all patients (NNT =27) and 39% (NNT=23) when excluding patients admitted on the same day as treatment (Table 2). For study subjects treated within five days of symptom onset, the hospitalization RRR was 40%, (NNT=17), even when including subjects hospitalized on the day of transfusion. We also found a 40% hospitalization RRR (NNT=21) for patients that received CCP with SARS-CoV2 antibody titers above the median level for each individual study. Early treatment and high antibody levels indicated a 51% hospitalization RRR (NNT=13). Despite differences in the demographics and clinical characteristics of the five study populations, overall study heterogeneity was low to moderate, suggesting the appropriateness of combining these studies in a single meta-analysis and broadly generalizing these results. While the effectiveness of early CCP treatment in reducing all-cause hospitalization was less than that of many monoclonal antibody treatments^31, 32^ and antiviral therapies,^12, 33^ this should be balanced against its increased availability and potential for activity against variant strains of SARS-CoV-2.

Two of the five RCTs included in this meta-analysis (CONV-ERT and C3PO) failed to demonstrate a reduction in all-cause hospitalization with CCP, while the other three trials all showed approximately 50% reductions in hospitalizations (CCP-Argentina, CSSC-004, CoV-Early). One potential explanation for the lack of effectiveness for CCP in the CONV-ERT trial is that methylene blue photo inactivation was used for pathogen reduction in transfused units. This might have affected the constant regions of antibody function without interfering with the viral neutralization assay.^34^ Importantly, this method of pathogen reduction is not used in the US. The C3PO trial, unlike the other RCTs, enrolled only patients presenting to the Emergency Department (ED) with COVID-19, which likely included a more severely ill patient population further in the inflammatory phase even after controlling for the days of symptoms, demographic, and identified risk factors. Indeed, there are often less tangible factors signifying more severe illness that lead a patient to present to the ED rather than to their primary care doctor. This is evidenced by the much larger number of subjects in the C3PO trial (23% of all hospitalizations) who were admitted directly to the hospital from the ED on the same visit in which they were transfused. This imbalance might be related to random chance, a difference in immediate reaction to CCP, or to the longer duration of observation required for patients in the CCP arm. Eliminating these same-day admissions (as in our secondary analysis) brings the C3PO results in line with those from the other studies and greatly reduces heterogeneity among the five studies.

Antibody levels for the transfused CCP used across these five trials varied substantially, despite the fact that donors had been selected based upon a minimum antibody level cut-off in each trial. However, different cut-offs were used as well as different antibody tests. Our observation that the effect on hospital admission was limited to patients receiving CCP with titers above the median concentration level in each of the trials suggests that the CCP selection process was suboptimal. It is likely that more stringent antibody titer criteria for CCP units may further improve the effectiveness of this intervention.^35^

Plasma transfusion, unlike the use of antiviral and monoclonal antibody agents, presents a risk of transfusion reactions, which may vary from easily treatable conditions (e.g., urticaria) to life-threatening reactions such as TACO, TRALI, and anaphylaxis. Rates of severe adverse reactions, however, appeared to be low in all of the included trials.

This study does have several important limitations. While CSSC-004 enrolled both COVID-19 vaccinated and unvaccinated individuals, the other RCTs primarily included unvaccinated patients, which limits our ability to analyze the effectiveness of CCP for reducing COVID-19 hospitalization in a primarily vaccinated population. The number needed to treat with CCP may be much higher in a primarily vaccinated population, although this difference may be mitigated by the rise of mutant variants that undermine the effectiveness of vaccines and mAbs.

Our meta-analysis chose to use a modified intention to treat analysis, excluding patients who were randomized to a given treatment but did not receive it which could introduce bias. However, this only affected a small number of patients, and would be unlikely to significantly affect our results. In one study, some patients not actually admitted to a hospital were considered to meet the primary outcome, but these patients did meet standard hospital admission criteria (i.e., hypoxia / respiratory distress) and were instead provided with hospital level care within their long-term care unit. As described above, the actual antibody titer levels varied across the five RCTs, and the studies used varying assays to measure antibody titer, making it difficult to compare absolute antibody titers across studies. Consequently, we chose to look at median antibody titers within the individual studies as a means of comparing the CCP used in the various RCTs.

Although there are several implementation considerations that could affect the real-world efficacy and sustainability of CCP transfusion programs^36^, our pooled meta-analysis including five large, rigorously conducted RCTs suggests that high-titer CCP administered early to adult outpatients with COVID-19 significantly reduces the risk of all-cause hospitalizations across a diverse range of demographic and clinical profiles, geographic locations, and transfusion settings. We believe that CCP should be considered as an outpatient treatment option (especially for patients at high-risk for poor outcomes) in settings where monoclonal antibodies or antivirals are not currently accessible, or when new variants arise undermining the effectiveness of these interventions. Future research should focus on defining the optimal antibody titer and dosage for CCP, and evaluating its effectiveness among immunocompromised vaccinated patients. Despite its limitations, CCP has the potential to be an effective, readily available, and highly adaptable intervention for use in both this and future pandemics.

## Data Availability

Data is available from individual authors upon request.

## Acknowledgements

We thank the patients that participated and all the plasma donors as well as all researchers and study nurses involved at the study sites.

## Authors’ contributions

Drs. Levine and Sullivan had full access to all the data in the study and takes responsibility for the integrity of the data and the accuracy of the data analysis

Concept and design: Sullivan, Hanley, Levine

Acquisition, analysis, or interpretation of data: Levine, Fukuta, Huaman, Ou, Meisenberg, Patel, Paxton, Hanley, Rijnders, Gharbharan, Rokx, Zwaginga, Alemany, Mitjà, Durkalski-Mauldin, Dumont, Korley, Callaway, Libster, Sullivan.

Drafting of the manuscript: Sullivan, Levine, Fukuta, Huaman, Ou, Meisenberg, Patel, Paxton. Critical revision of the manuscript for important intellectual content: Levine, Fukuta, Huaman, Ou,

Meisenberg, Patel, Paxton. Hanley, Rijnders, Gharbharan, Alemany, Mitjà, Durkalski-Mauldin, Korley, Callaway, Libster, Sullivan, Rokx, Zwafinga, Millat-Martinez.

Statistical analysis: Ou, Sullivan, Levine

Obtained funding: Sullivan, Rijinders, Mitja, Callaway, Polack.

Administrative, technical, or material support: Rokx, Zwaginga, Millat-Martinez.

Supervision: Sullivan, Gharbharan, Alemany, Callaway, Libster

## Data Sharing Statement

Data is available from individual authors upon request.

## Figure Legends

**Figure 1**. PRISMA chart. The MEDLINE, Embase, MedRxiv, Cochrane Library, WHO COVID-19 Research Database, and Web of Science were searched for all RCTs as of 30 September 2022.

**Figure 2**. Forest plot of A) modified Intention to Treat Analysis and B) of modified Intention to Treat Analysis excluding same day hospital admissions

**Figure 3**. Forest plots of transfusion A) within 5 days or B) greater than 5 days

**Figure 4**. Forest plots plasma donor antibody levels A) at or above median titer or B) less than median titer

**Figure 5**. Forest plots plasma donor antibody levels and early treatment A) at or above median titer AND transfusion within 5 days or B) Total of (Low titer and onset<= 5 days, High titer and onset over 5 days, Low titer and onset over 5 days

Supplementary material for Levine, Fukata …

**Supplementary Table 1**. Risk of bias

**Supplementary Table 2**. COVID-19 convalescent plasma characterization

**Supplementary Figure 1** Literature search strategies

**Supplementary Figure 2**. Funnel plot analysis

**Supplementary Figure 3**. Viral neutralization by study

**Supplementary Table 1.** Risk of bias

| **Risk of Bias** | **CSSC-004** | **CCP-Argentina** | **CONV-ERT** | **C3PO** | **\*CoV-Early** |
| --- | --- | --- | --- | --- | --- |
| Randomization | Low | Low | Low | Low | Some concerns |
| Deviations from intervention | Low | Low | Low | Low | Low |
| Missing outcome data | Low | Low | Low | Low | Low |
| Measurement of the outcome | Low | Low | Low | Low | Low |
| Selection of the reported results | Low | Some concerns | Low | Low | Low |
| Overall risk of bias | Low | Some concerns | Low | Low | Some concerns |

\*COVID-NMA ran ROB on compiled real time pooling analysis of only CONV-ERT and CoV-Early studies CONV-ERT and CoV-Early with some concerns for CoV-Early “No information on allocation concealment”.

**Supplementary Table 2.**
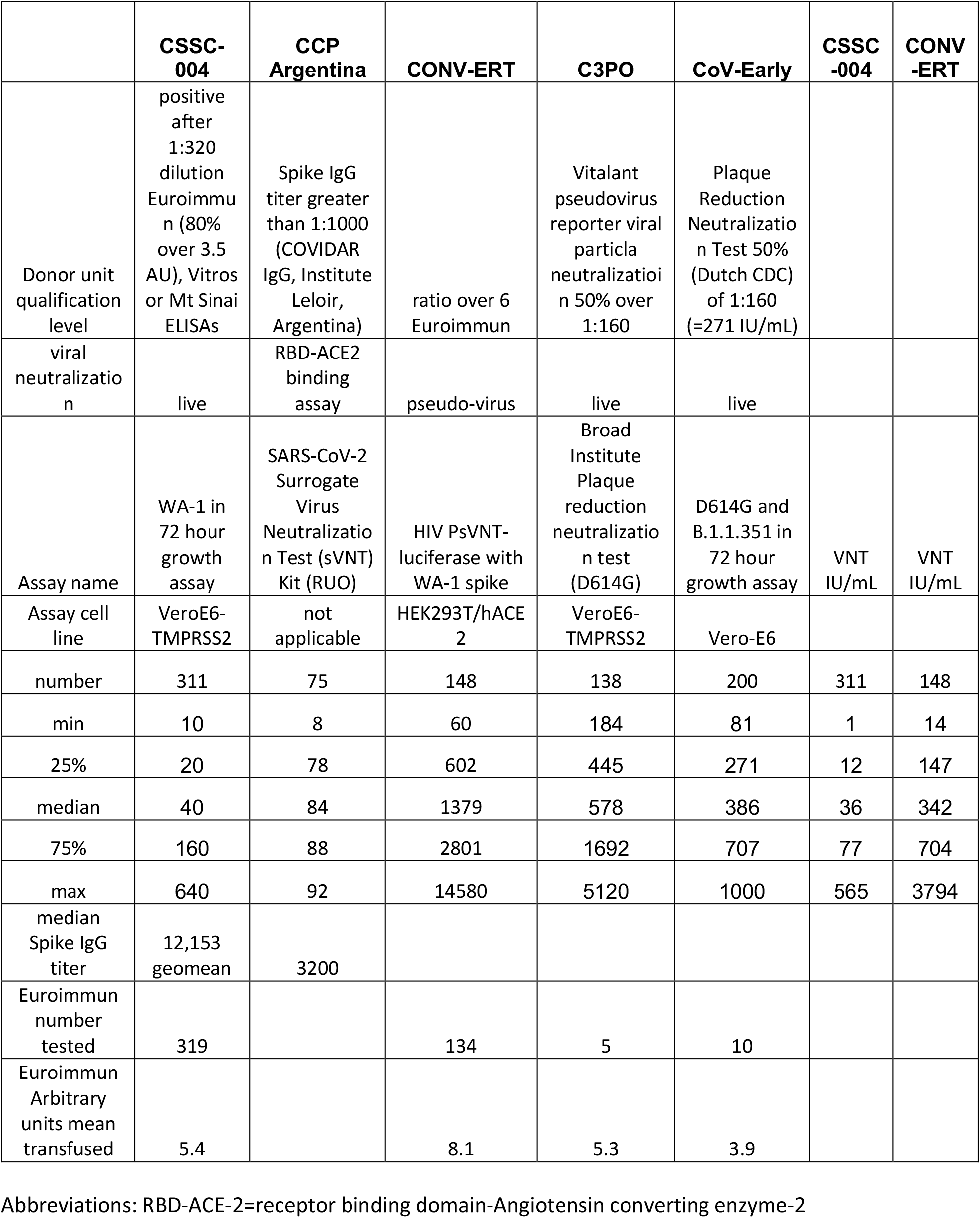
COVID-19 convalescent plasma characterization CCP-Argentina remnant donor plasma not available for Euroimmun testing

**Supplementary Fig 1.**
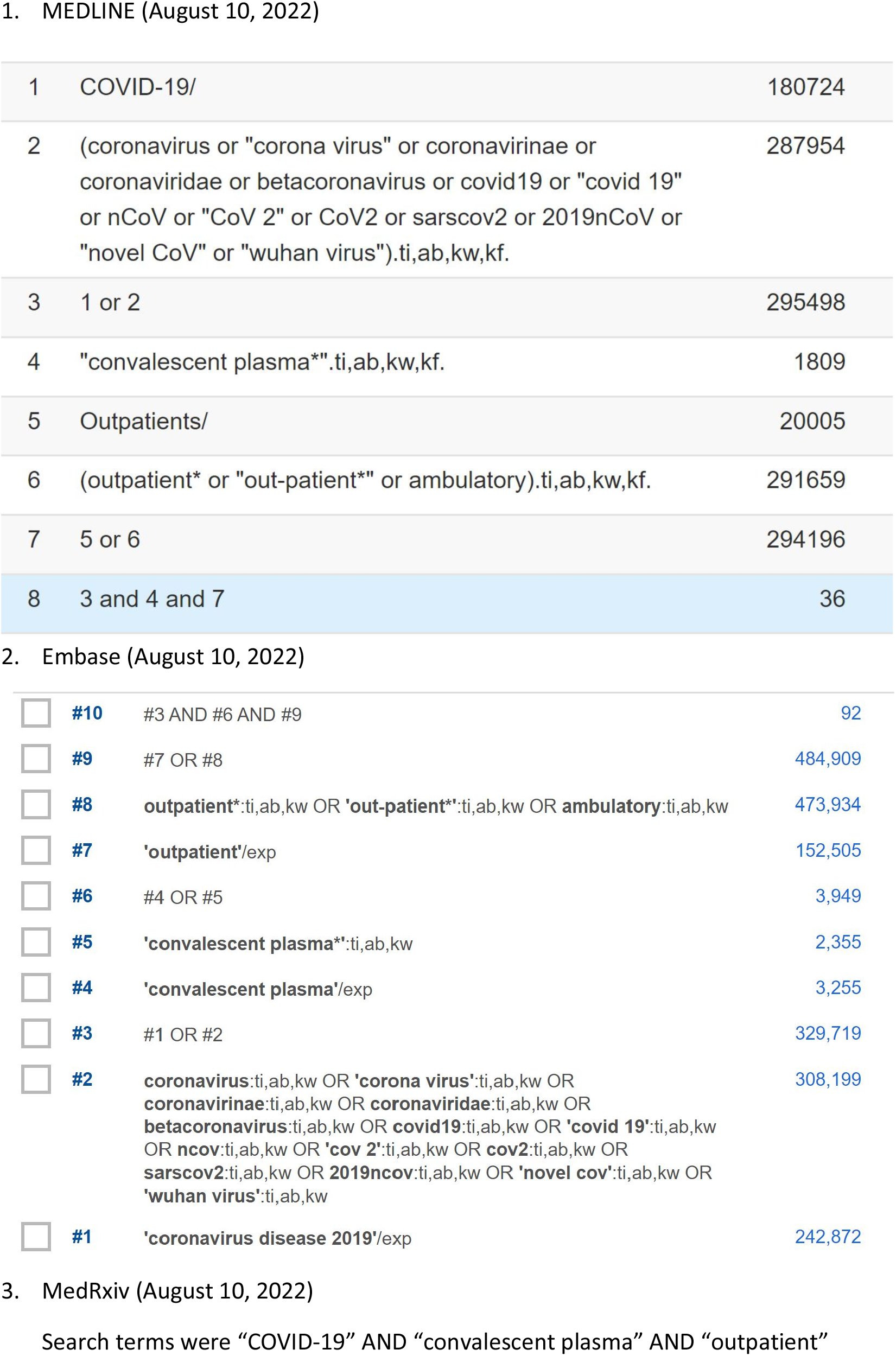
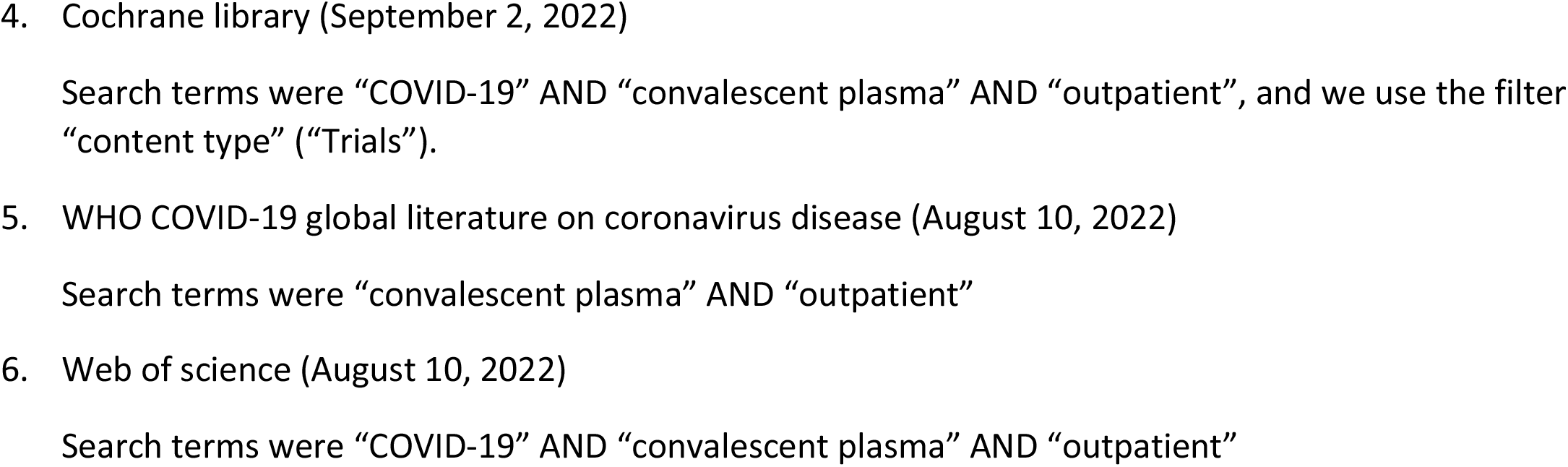
Literature search strategies

**Supplementary Figure 2.**
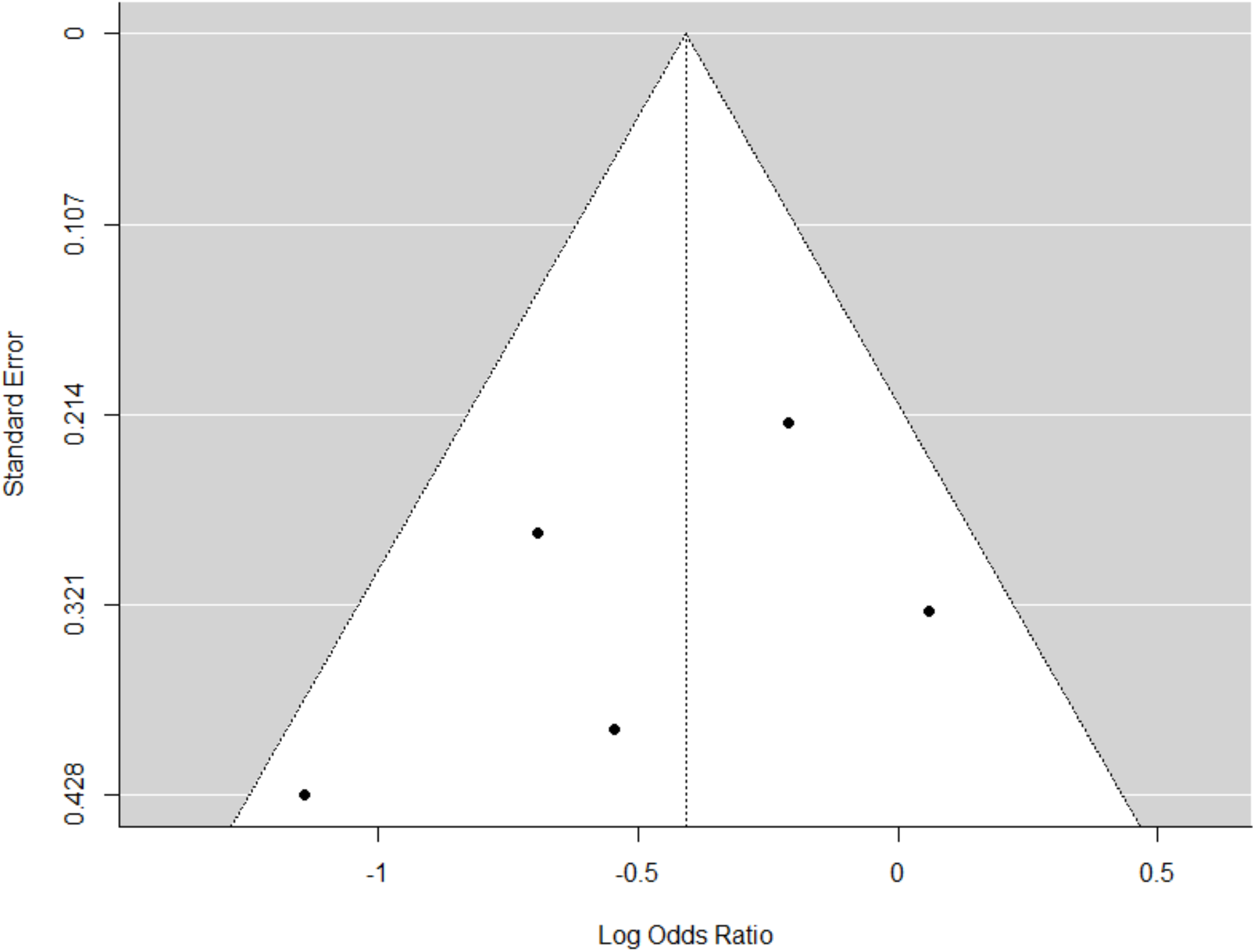
Funnel plot analysis

**Supplementary Figure 3.**
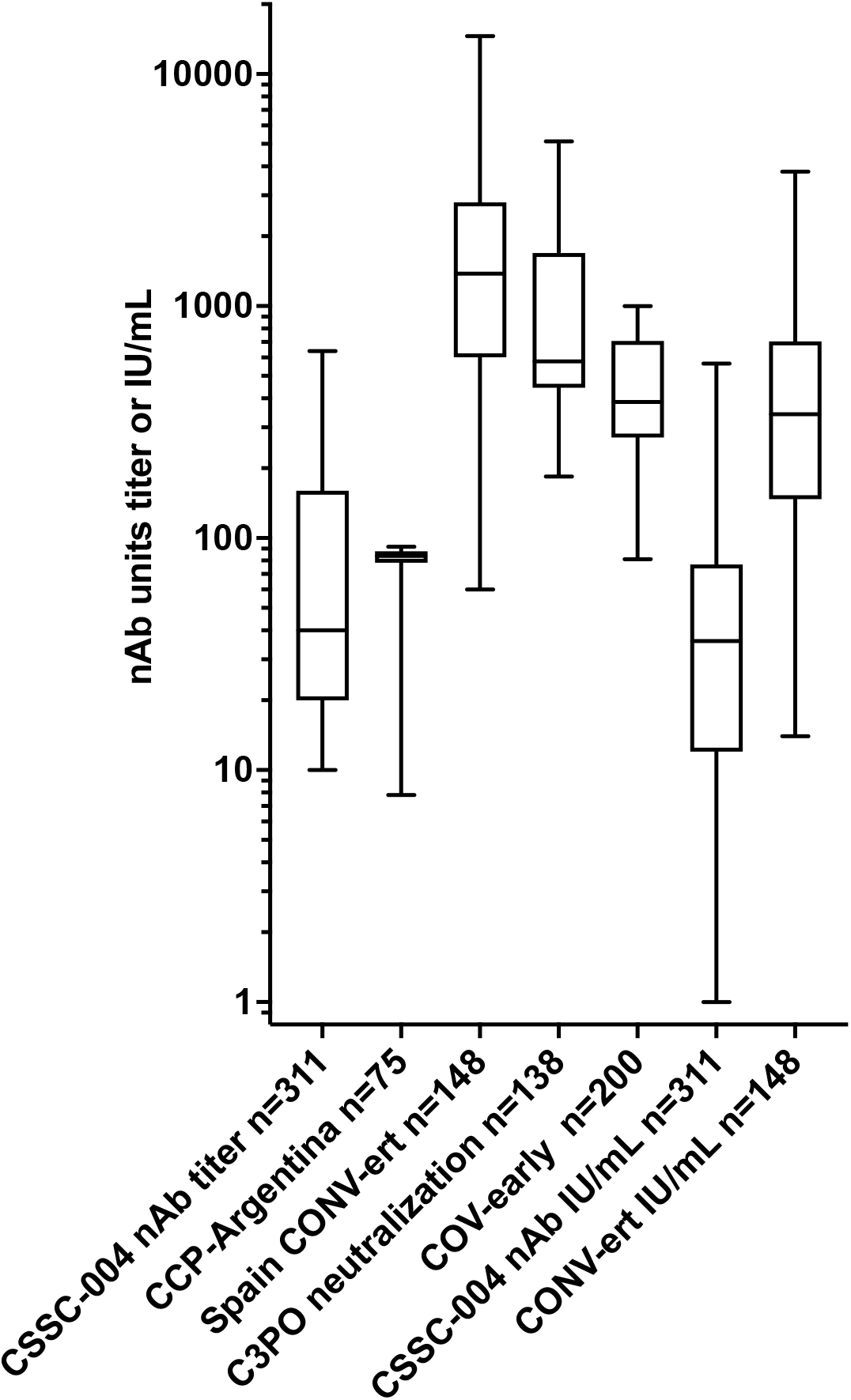
Viral neutralization by study. CSSC-004 used WA-1 virus in VeroE6-TMPRSS2 cells, CCP-Argentina used an RBD to ACE2 binding assay, CONV-ERT utilized an HIV pseudovirus in HEK293T/hACE2 cells with WA-1 spike, C3PO used D614G virus in VeroE6-TMPRSS2 cells, and CoV-Early used D614G and B1.12.351 in Vero-E6 cells with dilutional titers to interfere graphed. CSSC-004 in this depiction of diverse tests appeared to be lower than the other studies in range of viral neutralization. The minimum, 25%, median, 75% and maximum dilutional titers to interfere with 50% of assay signal is shown.

## References

1. Collaborators C-EM. Estimating excess mortality due to the COVID-19 pandemic: a systematic analysis of COVID-19-related mortality, 2020-21. Lancet. Apr 16 2022;399(10334):1513–1536. doi:10.1016/S0140-6736(21)02796-3

2. Paredes MI, Lunn SM, Famulare M, et al. Associations between SARS-CoV-2 variants and risk of COVID-19 hospitalization among confirmed cases in Washington State: a retrospective cohort study. medRxiv. Feb 16 2022;doi:10.1101/2021.09.29.21264272

3. Newland M, Durham D, Asher J, et al. Improving pandemic preparedness through better, faster influenza vaccines. Expert Rev Vaccines. Mar 2021;20(3):235–242. doi:10.1080/14760584.2021.1886931

4. Kachali H, Haavisto I, Leskela RL, Valja A, Nuutinen M. Are preparedness indices reflective of pandemic preparedness? A COVID-19 reality check. Int J Disaster Risk Reduct. Jul 2022;77:103074. doi:10.1016/j.ijdrr.2022.103074

5. Ramachandran R, Ross JS, Miller JE. Access to COVID-19 Vaccines in High-, Middle-, and Low-Income Countries Hosting Clinical Trials. JAMA Netw Open. Nov 1 2021;4(11):e2134233. doi:10.1001/jamanetworkopen.2021.34233

6. Salazar E, Christensen PA, Graviss EA, et al. Treatment of Coronavirus Disease 2019 Patients with Convalescent Plasma Reveals a Signal of Significantly Decreased Mortality. Am J Pathol. Nov 2020;190(11):2290–2303. doi:10.1016/j.ajpath.2020.08.001

7. Chen P, Nirula A, Heller B, et al. SARS-CoV-2 Neutralizing Antibody LY-CoV555 in Outpatients with Covid-19. N Engl J Med. Jan 21 2021;384(3):229–237. doi:10.1056/NEJMoa2029849

8. Weinreich DM, Sivapalasingam S, Norton T, et al. REGN-COV2, a Neutralizing Antibody Cocktail, in Outpatients with Covid-19. N Engl J Med. Jan 21 2021;384(3):238–251. doi:10.1056/NEJMoa2035002

9. Polack FP, Thomas SJ, Kitchin N, et al. Safety and Efficacy of the BNT162b2 mRNA Covid-19 Vaccine. N Engl J Med. Dec 31 2020;383(27):2603–2615. doi:10.1056/NEJMoa2034577

10. Anderson EJ, Rouphael NG, Widge AT, et al. Safety and Immunogenicity of SARS-CoV-2 mRNA-1273 Vaccine in Older Adults. N Engl J Med. Dec 17 2020;383(25):2427–2438. doi:10.1056/NEJMoa2028436

11. Weekly epidemiological update - 29 December 2020 (WHO) (2020).

12. Hammond J, Leister-Tebbe H, Gardner A, et al. Oral Nirmatrelvir for High-Risk, Nonhospitalized Adults with Covid-19. N Engl J Med. Apr 14 2022;386(15):1397–1408. doi:10.1056/NEJMoa2118542

13. Plata GG. The black market for covid-19 antiviral drugs. BMJ. May 31 2022;377:o1282. doi:10.1136/bmj.o1282

14. Hill A, Ellis L, Wang J, Pepperrell T. Prices versus costs of production for molnupiravir as a COVID-19 treatment. Research Square; 2021.

15. Dougan M, Azizad M, Chen P, et al. Bebtelovimab, alone or together with bamlanivimab and etesevimab, as a broadly neutralizing monoclonal antibody treatment for mild to moderate, ambulatory COVID-19. medRxiv. 2022:2022.03.10.22272100. doi:10.1101/2022.03.10.22272100

16. Levin MJ, Ustianowski A, De Wit S, et al. Intramuscular AZD7442 (Tixagevimab-Cilgavimab) for Prevention of Covid-19. N Engl J Med. Jun 9 2022;386(23):2188–2200. doi:10.1056/NEJMoa2116620

17. Vellas C, Kamar N, Izopet J. Resistance mutations in SARS-CoV-2 omicron variant after tixagevimab-cilgavimab treatment. J Infect. Jul 22 2022;doi:10.1016/j.jinf.2022.07.014

18. Huygens S, Munnink BO, Gharbharan A, Koopmans M, Rijnders B. Sotrovimab resistance and viral persistence after treatment of immunocompromised patients infected with the SARS-CoV-2 Omicron variant. Clin Infect Dis. Jul 22 2022;doi:10.1093/cid/ciac601

19. Birnie E, Biemond JJ, Appelman B, et al. Development of Resistance-Associated Mutations After Sotrovimab Administration in High-risk Individuals Infected With the SARS-CoV-2 Omicron Variant. Jama. Sep 20 2022;328(11):1104–1107. doi:10.1001/jama.2022.13854

20. Hernandez AV, Piscoya A, Pasupuleti V, et al. Beneficial and harmful effects of monoclonal antibodies for the treatment and prophylaxis of COVID-19: a systematic review and meta-analysis of randomized controlled trials. Am J Med. Jul 22 2022;doi:10.1016/j.amjmed.2022.06.019

21. Malahe SRK, Hoek RAS, Dalm V, et al. Clinical characteristics and outcome of immunocompromised patients with COVID-19 caused by the Omicron variant: a prospective observational study. Clin Infect Dis. Jul 23 2022;doi:10.1093/cid/ciac571

22. Bhimraj A MR, Shumaker AH, Baden L, Cheng VC, Edwards KM, Gallagher JC, Gandhi RT, Muller WJ, Nakamura MM, O’Horo JC, Shafer RW, Shoham S, Murad MH, Mustafa RA, Sultan S, Falck-Ytter Y. Infectious Diseases Society of America Guidelines on the Treatment and Management of Patients with COVID-19. Accessed September 30, 2022, https://www.idsociety.org/practice-guideline/covid-19-guideline-treatment-and-management/.

23. Page MJ, McKenzie JE, Bossuyt PM, et al. The PRISMA 2020 statement: an updated guideline for reporting systematic reviews. BMJ. Mar 29 2021;372:n71. doi:10.1136/bmj.n71

24. Boutron I, Chaimani A, Meerpohl JJ, et al. The COVID-NMA Project: Building an Evidence Ecosystem for the COVID-19 Pandemic. Ann Intern Med. Dec 15 2020;173(12):1015–1017. doi:10.7326/M20-5261

25. Nguyen TV, Ferrand G, Cohen-Boulakia S, et al. RCT studies on preventive measures and treatments for COVID-19. Zenodo. April 1, 2020 2020;doi:10.5281/zenodo.4266529

26. Sullivan DJ, Gebo KA, Shoham S, et al. Early Outpatient Treatment for Covid-19 with Convalescent Plasma. N Engl J Med. Mar 30 2022;doi:10.1056/NEJMoa2119657

27. Korley FK, Durkalski-Mauldin V, Yeatts SD, et al. Early Convalescent Plasma for High-Risk Outpatients with Covid-19. N Engl J Med. Nov 18 2021;385(21):1951–1960. doi:10.1056/NEJMoa2103784

28. Gharbharan A, Jordans C, Zwaginga L, et al. Outpatient convalescent plasma therapy for high-risk patients with early COVID-19. A randomized placebo-controlled trial. Clin Microbiol Infect. Aug 22 2022;doi:10.1016/j.cmi.2022.08.005

29. Alemany A, Millat-Martinez P, Corbacho-Monne M, et al. High-titre methylene blue-treated convalescent plasma as an early treatment for outpatients with COVID-19: a randomised, placebo-controlled trial. Lancet Respir Med. Mar 2022;10(3):278–288. doi:10.1016/S2213-2600(21)00545-2

30. Libster R, Perez Marc G, Wappner D, et al. Early High-Titer Plasma Therapy to Prevent Severe Covid-19 in Older Adults. N Engl J Med. Feb 18 2021;384(7):610–618. doi:10.1056/NEJMoa2033700

31. Verderese JP, Stepanova M, Lam B, et al. Neutralizing Monoclonal Antibody Treatment Reduces Hospitalization for Mild and Moderate Coronavirus Disease 2019 (COVID-19): A Real-World Experience. Clin Infect Dis. Mar 23 2022;74(6):1063–1069. doi:10.1093/cid/ciab579

32. Jenks JD, Aslam S, Horton LE, et al. Early Monoclonal Antibody Administration Can Reduce Both Hospitalizations and Mortality in High-Risk Outpatients With Coronavirus Disease 2019 (COVID-19). Clin Infect Dis. Mar 1 2022;74(4):752–753. doi:10.1093/cid/ciab522

33. Jayk Bernal A, Gomes da Silva MM, Musungaie DB, et al. Molnupiravir for Oral Treatment of Covid-19 in Nonhospitalized Patients. N Engl J Med. Feb 10 2022;386(6):509–520. doi:10.1056/NEJMoa2116044

34. Ross V. Photodynamic action of methylene blue on diphtheric antitoxin. J Immunol. Nov 1938;35(5):371–377.

35. Rijnders BJA, Huygens S, Mitja O. Evidence-based dosing of convalescent plasma for COVID-19 in future trials. Clin Microbiol Infect. May 2022;28(5):667–671. doi:10.1016/j.cmi.2022.01.026

36. Bloch EM, Tobian AAR, Shoham S, et al. How do I implement an outpatient program for the administration of convalescent plasma for COVID-19? Transfusion. May 2022;62(5):933–941. doi:10.1111/trf.16871

